# The Cartilage Thickness Score (CTh-Score) Captures High-Resolution Cartilage Thickness Patterns Associated with Osteoarthritis Onset, Progression, and Knee Replacement: Data from the Osteoarthritis Initiative

**DOI:** 10.64898/2026.04.14.26350838

**Authors:** Paul Margain, Julien Favre, C. Kent Kwoh, Patrick Omoumi

## Abstract

**Objective:** To evaluate the Cartilage Thickness Score (CTh-Score) as an MRI-based quantitative measure of cartilage damage severity by assessing its association with osteoarthritis (OA) clinical course milestones and comparing its performance with conventional morphometric measures, including radiographic minimum joint space width (JSW) and regional average cartilage thickness.

**Methods:** Three matched case-control analyses from the Osteoarthritis Initiative included 1,669 knees: incident radiographic OA (304 cases/304 controls), pain and structural progression (164/369), and knee replacement (KR) within 2 years (132/396). CTh-Score and conventional morphometric measures were calculated at 4 years (T_−4Y_), 2 years (T_−2Y_), and T_0_ before each milestone. Analyses evaluated between-group differences, longitudinal change, responsiveness, and associations independent of demographics and radiographic severity.

**Results:** Across all study designs, CTh-Score differentiated cases from controls at every timepoint. In the OA onset cohort, cases had higher CTh-Scores than controls at T_−4Y_ (16.0 vs 12.1, p=0.002), T_−2Y_ (23.4 vs 14.9, p<0.001), and T_0_ (39.7 vs 17.3, p<0.001; n=304/304). Conventional morphometric measures showed weaker or later discrimination. Similar patterns were observed for progression and KR cohorts. Longitudinal CTh-Score changes distinguished cases from controls earlier and more consistently, with greater responsiveness than conventional measures. CTh-Score was the only measure independently associated with all milestones in adjusted models at T_−4Y_ and T_−2Y_.

**Conclusion:** The CTh-Score captures high-resolution cartilage thickness patterns associated with OA onset, progression, and KR, outperforming conventional morphometric measures in early discrimination, responsiveness, and predictive association. These findings support CTh-Score as a sensitive quantitative marker of cartilage damage severity across the OA clinical course continuum.

## Introduction

Although knee osteoarthritis (OA) is now recognized as a whole-organ disease affecting all joint structures [1], articular cartilage degeneration remains a central hallmark of the disease. Cartilage deterioration progresses across all disease stages and encompasses a broad spectrum of structural changes, ranging from early swelling and surface fibrillation to ultimately full-thickness cartilage loss [2].

Cartilage thickness forms distinct spatial patterns that can be captured at high-spatial-resolution using MRI [3–5]. These patterns carry meaningful information about OA severity, as shown by associations with disease severity in previous studies [4, 6]. However, high-resolution spatial patterns of cartilage thickness remain underexplored due to two main methodological challenges. First, achieving accurate spatial correspondence of subchondral bone surfaces across individuals is technically complex, making direct pattern comparison difficult. Second, the high inter-individual variability of spatial patterns complicates their interpretation using conventional approaches. To circumvent these challenges, traditional quantitative assessments of cartilage morphology rely on indirect surrogates such as minimum joint space width on radiographs [7], or predefined regions of interest (ROIs), measuring average cartilage thickness on MRI [8]. Although these low-spatial-resolution metrics have demonstrated extensive clinical utility, particularly in longitudinal studies where lesion locations are more consistent within individuals, they capture only a partial amount of the spatial information content that is available from the underlying cartilage thickness patterns.

Recently, the methodological barriers preventing the assessment of spatial patterns of cartilage thickness have been addressed through the introduction of an automated framework that generates anatomically standardized, high-resolution femorotibial cartilage thickness maps (CTh-Maps) [5]. One straightforward way to interpret the significant wealth of information in these maps is the Cartilage Thickness Score (CTh-Score). This continuous severity metric summarizes spatial cartilage thickness patterns on a standardized 0–100 scale, ranging from healthy cartilage to end-stage OA. This score has demonstrated high reproducibility and validated against expert MRI-based assessments [5].

OA is typically slowly progressive [9], yet its clinical course can be described by key milestones that are directly meaningful to patients and healthcare systems: (i) the onset of structural disease as defined radiographically, a key component of the diagnosis, (ii) subsequent progression of symptoms and/or structural damage (which may not evolve concurrently), and (iii) knee replacement surgery marking end-stage disease. Semi-quantitative MRI-based assessments of cartilage damage [10, 11] have revealed associations with disease onset [12], progression [13], and future knee replacement [14].

The objective of this study was to evaluate the CTh-Score as a quantitative measure of cartilage damage severity by assessing its association with three clinically relevant milestones in the clinical course of knee OA and comparing its performance with conventional morphometric measures. Specifically, using three frequency matched case-control designs corresponding to the development of incident radiographic OA (ROA), combined pain and structural progression (from the FNIH Biomarkers Consortium cohort), and receipt of knee replacement, we assessed whether (a) the CTh-Score differentiates cases from controls up to four years before attainment of each milestone, (b) longitudinal changes in the CTh-Score differ between cases and controls, and (c) the CTh-Score demonstrates superior responsiveness and predictive ability compared with regional average cartilage thickness and minimum joint space width.

## Methods

### Ethics

The Osteoarthritis Initiative was approved by the institutional review boards of the participating centers, and all participants provided written informed consent. Participants were not involved in the design, conduct, reporting or dissemination plans of this secondary analysis.

### Study population and imaging acquisition

Data were obtained from the Osteoarthritis Initiative (OAI), a multi-center, longitudinal, prospective observational study of knee OA [15]. This study analyzed specific timepoints, including the baseline and follow-up visits up to 120 months, with annual in-person visits up to 48 months and then alternating annual telephone and in-person visits until 120 months. At each in person visit, participants underwent standardized bilateral fixed-flexion knee radiography. Bilateral MRI was acquired at in person visits up to 96 months using a standardized protocol on four identical Siemens 3T MRs, including the sagittal double-echo steady-state (DESS) sequence, which provides high-resolution 3D information suitable for cartilage morphometry [15].

### Image reading and quantitative measurements

Radiographic disease severity was assessed using the Kellgren-Lawrence (KL) grading system [16], derived from the OAI central reading repositories. Minimum joint space width (JSW) measurements, obtained using a previously validated semi-automated software, were retrieved from the OAI dataset [17].

For MRI-based cartilage assessment, high-resolution cartilage thickness maps (CTh-Maps) and the corresponding CTh-Score were obtained from a public repository [18]. To serve as a comparator for conventional morphometry, the average cartilage thickness was calculated within standard subregions of interest (medial and lateral femoral weight-bearing regions and tibial compartments) using previously validated methods [19–21].

### Study design

To evaluate the CTh-Score across the disease continuum, we employed three distinct matched case-control designs corresponding to key OA milestones: incident ROA, disease progression, and receipt of knee replacement as a clinical outcome of end-stage disease.

i. Incident ROA: cases were defined as knees transitioning from a non-OA state to established OA. Inclusion criteria required a KL grade <2 at two consecutive timepoints T_-4Y_ and T_-2Y_ followed by a conversion to KL ≥2 at the index visit T_0_. Cases were frequency matched to control knees that maintained a KL grade <2 throughout all timepoints (T_-4Y_, T_-2Y_, and T_0_). Matching was performed based on age, sex, BMI, baseline KL grade at T_-4Y_, and the specific OAI visits.
ii. Pain and structural progression: Data for this analysis were derived from the FNIH Biomarkers Consortium study, a nested case-control project within the OAI (n=600, one knee per participant) [22]. Progression was assessed at the 48-month visit (T_0_) relative to baseline (T_-4Y_). Pain progression was defined as an increase in WOMAC pain score ≥ 9 points, while structural progression was defined as a medial JSW loss ≥ 0.7 mm. We compared “pain and structural progressor” cases (i.e., had both pain and structural progression) against a pooled control group comprising three subsets: isolated pain progressors, isolated radiographic progressors, and non-progressors (i.e. no pain or structural progression). Groups were matched at baseline visit (T_-4Y_) for radiographic severity (including KL and JSW) and demographic factors (including use of analgesic and WOMAC pain).
iii. Knee replacement outcomes: Cases included participants who reported a knee replacement (KR) surgery during the study follow-up. The index time point (T_0_) was defined as the OAI visit closest to surgery with both MRI and radiographic data available, within a maximum window of 2 years before the operation. Cases were frequency matched to controls who did not undergo KR surgery during the study period. Matching was performed using data from two visits prior to the index date (T_-4Y_) based on age, sex, BMI, and KL grade. If a control participant had two eligible knees, the knee with the highest KL grade was selected.

### Statistical analysis

Analyses were performed using all available cases with complete data for the variables required in each model or comparison. All statistical tests were two-sided, and p values <0.05 were considered statistically significant.

Statistical analyses were performed using non-parametric tests because the distributions of CTh-Score were non-normal. To evaluate the sensitivity of the metrics to OA milestones, we compared CTh-Score, regional average thickness, and JSW between cases and controls at the matching baseline and subsequent follow-up visits using the Mann-Whitney U test.

The association of the CTh-Score was assessed using multivariable logistic regression models. Standardized odds ratios (OR) were calculated per standard deviation increase in the metric to predict case status (Onset, Progression, or KR). Models were adjusted for age, sex, BMI, and KL grade. For regional average thickness, values from all four tibiofemoral compartments were entered into a single model to determine independent contributions. JSW was excluded from the analysis of the Progression group as it was a defining criterion for the outcome.

Responsiveness was evaluated by calculating the longitudinal change in metrics over 2-year and 4-year intervals relative to the matching timepoint T_-4Y_. The Standardized Response Mean (SRM) was calculated as the mean change divided by the standard deviation of the change. The SRM was used to compare the responsiveness of the CTh-Score against regional averages and JSW.

## Results

### Associations with radiographic onset

A total of 304 knees with incident ROA (KL <2 at T_-4Y_ and T_-2Y_, progressing to KL ≥ 2 at T0) were matched 1:1 with control knees that remained KL <2 throughout follow-up. Matching was successful for age, sex, BMI, and baseline KL grade (Table 1).

**Table 1.** Case–control comparison of cartilage morphology before incident radiographic osteoarthritis (onset cohort). Cross-sectional and longitudinal comparisons between incident radiographic OA cases (KL < 2 at T_−4Y_ and T_−2Y_, converting to KL ≥ 2 at T_0_) and matched controls remaining KL < 2 at all timepoints. Values are median (IQR) for continuous variables and percentages for categorical variables. P values compare cases vs controls at each timepoint and for change scores (Δ). Timepoints: T_0_, index visit at radiographic onset (KL ≥ 2); T_−2Y_ and T_−4Y_, 2 and 4 years before T_0_. Measures: CTh-Score (0–100), medial minimum JSW (mm), and ROI-averaged cartilage thickness (mm) for femoral weight-bearing medial, femoral weight-bearing lateral, tibial medial, and tibial lateral compartments.

|  |  | Case<sub>Onset</sub> | Control<sub>Onset</sub> | p |
| --- | --- | --- | --- | --- |
|  | Number of knees | 304 | 304 |  |
| T<sub>-4Y</sub> | **Matched variables** |  |  |  |
|  | Age (y) | 61.0 (55.8;69.0) | 61.0 (56.0;69.0) | 0.90 |
|  | Sex (%F) | 65.80% | 65.80% | 1 |
|  | BMI (kg/m<sup>2</sup>) | 28.0 (25.3;31.3) | 27.8 (25.1;31.5) | 0.83 |
|  | KL0/KL1 (%) | 41.1/58.9 | 41.1/58.9 | 1 |
|  | **Non-matched variables** |  |  |  |
|  | JSW (mm) | 4.5 (4.0;5.1) | 4.4 (3.9;4.9) | 0.13 |
|  | Femur medial (mm) | 2.2 (2.0;2.4) | 2.2 (2.0;2.4) | 0.88 |
|  | Femur lateral (mm) | 2.3 (2.2;2.5) | 2.3 (2.2;2.5) | 0.99 |
|  | Tibia medial (mm) | 1.7 (1.6;1.9) | 1.7 (1.6;1.9) | 0.90 |
|  | Tibia lateral (mm) | 2.0 (1.8;2.2) | 2.0 (1.8;2.2) | 0.16 |
|  | CTh-Score (0-100) | 16.0 (8.1;24.4) | 12.1 (5.8;20.4) | **0.002** |
| T<sub>-2Y</sub> | JSW | 4.4 (3.8;4.9) | 4.3 (3.9;4.8) | 0.08 |
|  | Femur medial | 2.2 (2.0;2.4) | 2.2 (2.0;2.4) | 0.81 |
|  | Femur lateral | 2.3 (2.1;2.5) | 2.3 (2.1;2.5) | 0.32 |
|  | Tibia medial | 1.7 (1.6;1.9) | 1.7 (1.5;1.9) | 0.71 |
|  | Tibia lateral | 1.9 (1.7;2.2) | 2.0 (1.8;2.2) | 0.40 |
|  | CTh-Score | 23.4 (14.7;32.6) | 14.9 (7.4;25.1) | **<0.001** |
| T<sub>0</sub> | JSW | 3.8 (2.9;4.6) | 4.2 (3.8;4.8) | **<0.001** |
|  | Femur medial | 2.1 (1.9;2.3) | 2.2 (1.9;2.4) | 0.06 |
|  | Femur lateral | 2.3 (2.1;2.5) | 2.3 (2.1;2.5) | 0.77 |
|  | Tibia medial | 1.7 (1.5;1.9) | 1.7 (1.5;1.9) | 0.84 |
|  | Tibia lateral | 1.9 (1.6;2.2) | 1.9 (1.7;2.2) | 0.09 |
|  | CTh-Score | 39.7 (27.1;49.6) | 17.3 (7.9;29.1) | **<0.001** |
| T<sub>-2Y</sub> - T<sub>-4Y</sub> | ΔJSW | -0.12 (-0.46;0.12) | -0.05 (-0.27;0.17) | **0.003** |
|  | ΔFemur medial | -0.02 (-0.08;0.05) | -0.01 (-0.08;0.05) | 0.08 |
|  | ΔFemur lateral | -0.00 (-0.04;0.05) | -0.01 (-0.05;0.03) | **0.003** |
|  | ΔTibia medial | -0.01 (-0.06;0.03) | -0.02 (-0.07;0.02) | 0.23 |
|  | ΔTibia lateral | -0.03 (-0.08;0.02) | -0.04 (-0.08;0.01) | 0.07 |
|  | ΔCTh-Score | 5.1 (1.3;9.7) | 1.8 (-0.2;5.0) | **<0.001** |
| T<sub>0</sub> - T<sub>-4Y</sub> | ΔJSW | -0.63 (-1.4;0.00) | -0.15 (-0.42;0.08) | **<0.001** |
|  | ΔFemur medial | -0.03 (-0.17;0.05) | -0.02 (-0.10;0.04) | **0.05** |
|  | ΔFemur lateral | -0.01 (-0.08;0.05) | -0.02 (-0.07;0.02) | 0.27 |
|  | ΔTibia medial | -0.04 (-0.09;0.04) | -0.03 (-0.08;0.02) | 0.84 |
|  | ΔTibia lateral | -0.06 (-0.14; -0.01) | -0.07 (-0.13; -0.02) | 0.96 |
|  | ΔCTh-Score | 18.7 (10.2;31.5) | 3.0 (0.48;7.8) | **<0.001** |

At T_-4Y_ (4 years prior to incident ROA), the CTh-Score was already significantly higher in case knees than in controls (median [IQR]: 16.0 [8.1; 24.4] vs. 12.1 [5.8; 20.4]; p = 0.002), whereas JSW and all regional mean cartilage thickness measures showed no significant between-group differences. This separation increased at T_-2Y_ (23.4 [14.7; 32.6] vs. 14.9 [7.4; 25.1]; p < 0.001) and became pronounced at T_0_ (39.7 [27.1; 49.6] vs. 17.3 [7.9; 29.1]; p < 0.001). At the time of onset, JSW (p < 0.001) also differed significantly between groups, while all regional mean cartilage thickness measures did not.

Longitudinally, changes in the CTh-Score differentiated cases from controls earlier and more consistently than conventional measures. Between T_-4Y_ and T_-2Y_, the increase in CTh-Score was greater in cases (Δ 5.1 [1.3; 9.7]) than controls (Δ 1.8 [-0.2; 5.0]; p < 0.001), despite minimal and inconsistent changes in regional thickness (Table 1). Over the full 4-year interval (T_-4Y_ to T_0_), cases exhibited a markedly larger increase in CTh-Score (Δ 18.7 [10.2; 31.5]) compared with controls (Δ 3.0 [0.48; 7.8]; p < 0.001), exceeding the magnitude of change observed for JSW and all ROI-based thickness metrics (SRM=1.41 vs SRM≤0.71 Supplementary Table 1).

In multivariable logistic regression adjusted for age, sex, BMI, and baseline KL grade (Table 2), the CTh-Score was independently associated with incident radiographic OA at both T_-4Y_ (OR per SD increase: 1.3 [1.1; 1.6], p = 0.002) and T_-2Y_ (1.7 [1.4; 2.1], p < 0.001). No regional average cartilage thickness measure demonstrated an independent association across these timepoints.

**Table 2.** Adjusted associations between cartilage morphology metrics and osteoarthritis milestones. Standardized odds ratios (ORs) per 1 SD increase in each metric for predicting case status in the three matched case–control designs (onset, progression, and KR) at T_−4Y_ and T_−2Y_. Models are adjusted for age, sex, BMI, and KL. For ROI-based cartilage thickness, femoral medial, femoral lateral, tibial medial, and tibial lateral measures are entered simultaneously in a single model to estimate independent contributions. Medial minimum JSW is not reported for the progression analysis because JSW loss defines the structural progression outcome.

|  | Onset |  | Progression |  | KR |  |
| --- | --- | --- | --- | --- | --- | --- |
|  | T<sub>-4Y</sub> | T<sub>-2Y</sub> | T<sub>-4Y</sub> | T<sub>-2Y</sub> | T<sub>-4Y</sub> | T<sub>-2Y</sub> |
| JSW | 1.1<br>(0.94;1.3)<br>p=0.13 | 1.0<br>(0.80;1.1)<br>p=0.78 |  |  | 0.97<br>(0.76;1.2)<br>p=0.77 | 0.92<br>(0.73;1.2)<br>p=0.52 |
| Femur medial | 0.98<br>(0.81;1.2)<br>p=0.89 | 0.96<br>(0.79;1.2)<br>p=0.81 | 0.87<br>(0.71;1.1)<br>p=0.19 | **0.75**<br>**(0.61;0.91)**<br>**p<0.001** | 0.86<br>(0.68;1.1)<br>p=0.19 | 0.85<br>(0.67;1.1)<br>p=0.16 |
| Femur lateral | 1.1<br>(0.89;1.3)<br>p=0.99 | 1.1<br>(0.95;1.4)<br>p=0.32 | 1.1<br>(0.94;1.4)<br>p=0.18 | 1.2<br>(0.97;1.4)<br>p=0.11 | 1.05<br>(0.84;1.3)<br>p=0.67 | 1.04<br>(0.84;1.3)<br>p=0.71 |
| Tibia medial | 1.0<br>(0.83;1.2)<br>p=0.91 | 1.0<br>(0.85;1.2)<br>p=0.71 | 1.2<br>(1.0;1.5)<br>p=0.05 | 1.1<br>(0.91;1.4)<br>p=0.30 | 1.0<br>(0.79;1.3)<br>p=0.99 | 1.0<br>(0.79;1.2)<br>p=0.92 |
| Tibia lateral | 0.89<br>(0.76;1.1)<br>p=0.16 | 0.92<br>(0.77;1.1)<br>p=0.40 | 0.98<br>(0.79;1.2)<br>p=0.88 | 1.0<br>(0.83;1.27)<br>p=0.84 | 1.0<br>(0.76;1.2)<br>p=0.74 | 1.0<br>(0.79;1.3)<br>p=0.99 |
| CTh-Score | **1.3**<br>**(1.1;1.6)**<br>**p=0.002** | **1.7**<br>**(1.4;2.1)**<br>**p<0.001** | **1.5**<br>**(1.3;1.9)**<br>**p<0.001** | **1.8**<br>**(1.5;2.3)**<br>**p<0.001** | **2.0**<br>**(1.4;2.8)**<br>**p<0.001** | **1.9**<br>**(1.3;2.9)**<br>**p<0.001** |

### Associations with pain and structural progression

The progression analysis included 164 knees with combined pain and structural progression and 369 matched control knees from the FNIH cohort (Table 3). Groups were comparable at baseline with respect to age, sex, BMI, and KL grade distribution.

**Table 3.** Case–control comparison of cartilage morphology before combined pain and structural progression (FNIH cohort). Cross-sectional and longitudinal comparisons between knees with combined pain and structural progression (“progressors”) and matched controls from the FNIH Biomarkers Consortium nested case–control study. Values are median (IQR) for continuous variables and percentage for categorical variables. P values compare progressors vs controls at each timepoint and for change scores (Δ). JSW is shown at baseline but not reported longitudinally because JSW loss was part of the structural progression definition. Timepoints: T_0_ corresponds to the progression assessment visit; T_−2Y_ and T_−4Y_ denote 2 and 4 years before T_0_. Measures: CTh-Score (0–100), medial minimum JSW (mm), and ROI-averaged cartilage thickness (mm) for femoral weight-bearing medial, femoral weight-bearing lateral, tibial medial, and tibial lateral compartments.

|  |  | Case<sub>Progression</sub> | Control<sub>Progression</sub> | p |
| --- | --- | --- | --- | --- |
|  | Number of knees | 164 | 369 |  |
| T<sub>-4Y</sub> | **Matched variables** |  |  |  |
|  | Age (y) | 61.0 (54.0;69.0) | 61.0 (54;68.0) | 0.51 |
|  | Sex (%F) | 45.70% | 39.60% | 0.22 |
|  | BMI (kg/m<sup>2</sup>) | 30.4 (27.3;33.9) | 30.2 (27.6;33.6) | 0.78 |
|  | KL1/2/3(%) | 14.0/43.3/42.7 | 13.0/54.2/32.8 | 0.06 |
|  | JSW (mm) | 3.5 (2.8;4.74) | 3.9 (3.1;4.6) | 0.26 |
|  | **Non-matched variables** |  |  |  |
|  | Femur medial (mm) | 2.1 (1.9;2.4) | 2.2 (2.0;2.4) | 0.48 |
|  | Femur lateral(mm) | 2.4 (2.2;2.6) | 2.4 (2.2;2.6) | 0.3 |
|  | Tibia medial (mm) | 1.8 (1.6;2.0) | 1.7 (1.6;1.9) | 0.05 |
|  | Tibia lateral (mm) | 2.1 (1.8;2.3) | 2.0 (1.8;2.3) | 0.75 |
|  | CTh-Score (0-100) | 43.2 (24.3;57.9) | 33.0 (19.1;46.6) | **<0.001** |
| T<sub>-2Y</sub> | Femur medial | 2.1 (1.7;2.3) | 2.1 (1.9;2.4) | 0.06 |
|  | Femur lateral | 2.4 (2.2;2.6) | 2.3 (2.2;2.6) | 0.14 |
|  | Tibia medial | 1.8 (1.6;1.9) | 1.7 (1.6;1.9) | 0.39 |
|  | Tibia lateral | 2.0 (1.8;2.3) | 2.0 (1.8;2.3) | 0.51 |
|  | CTh-Score | 52.8 (33.3;65.7) | 37.9 (23.8;52.1) | **<0.001** |
| T<sub>0</sub> | Femur medial | 1.9 (1.5;2.2) | 2.1 (1.8;2.3) | **<0.001** |
|  | Femur lateral | 2.4 (2.2;2.5) | 2.3 (2.2;2.5) | 0.2 |
|  | Tibia medial | 1.7 (1.5;1.9) | 1.7 (1.5;1.9) | 0.48 |
|  | Tibia lateral | 2.0 (1.8;2.3) | 2.0 (1.8;2.2) | 0.66 |
|  | CTh-Score | 62.2 (47.8;73.9) | 42.3 (27.5;56.5) | **<0.001** |
| T<sub>-2Y</sub> - T<sub>-4Y</sub> | ΔFemur medial | -0.11 (-0.24; -0.01) | -0.04 (-0.14; 0.02) | **<0.001** |
|  | ΔFemur lateral | 0.00 (-0.06; 0.06) | 0.02 (-0.07; 0.04) | 0.13 |
|  | ΔTibia medial | -0.04 (-0.10; -0.01) | -0.02 (-0.07;0.03) | **<0.001** |
|  | ΔTibia lateral | -0.02 (-0.08; 0.04) | -0.03 (-0.10;0.02) | 0.13 |
|  | ΔCTh-Score | 7.6 (2.6;12.7) | 3.4 (0.33;8.4) | **<0.001** |
| T<sub>0</sub> - T<sub>-4Y</sub> | ΔFemur medial | -0.25 (-0.44; -0.07) | -0.06 (-0.20;0.03) | **<0.001** |
|  | ΔFemur lateral | -0.02 (-0.09; 0.06) | -0.02 (-0.07;0.04) | 1 |
|  | ΔTibia medial | -0.10 (-0.21; -0.01) | -0.04 (-0.10;0.01) | **<0.001** |
|  | ΔTibia lateral | -0.06 (-0.14; -0.01) | -0.06 (-0.11; -0.01) | 0.92 |
|  | ΔCTh-Score | 16.3 (10.4;27.6) | 6.8 (1.7;14.3) | **<0.001** |

At T_-4Y_, progressor knees already showed significantly higher CTh-Scores than controls (43.2 [24.3; 57.9] vs. 33.0 [19.1; 46.6]; p < 0.001). Regional cartilage thickness measures showed limited differences, with only tibial medial thickness reaching borderline significance. The separation in CTh-Score increased at T_-2Y_ (52.8 [33.3; 65.7] vs. 37.9 [23.8; 52.1]; p < 0.001) and further at T_0_ (62.2 [47.8; 73.9] vs. 42.3 [27.5; 56.5]; p < 0.001). At T_0_, medial femoral cartilage thickness was also significantly lower in progressors (p < 0.001), whereas other compartments were not (all p ≥ 0.2).

Longitudinal changes favored the CTh-Score as the most responsive metric (Supplementary Table 1). From T_-4Y_ to T_-2Y_, progressors demonstrated a greater increase in CTh-Score than controls (Δ 7.6 [2.6; 12.7] vs. 3.4 [0.33; 8.4]; p < 0.001). Over the full 4-year interval, the difference widened substantially (Δ 16.3 [10.4; 27.6] vs. 6.8 [1.7; 14.3]; p < 0.001), whereas changes in regional thickness were smaller and confined to medial compartments (Table 3).

In adjusted logistic regression models (Table 2), the CTh-Score was independently associated with progression status at both T_-4Y_ (OR per SD: 1.5 [1.3; 1.9], p < 0.001) and T_-2Y_ (1.8 [1.5; 2.3], p < 0.001). Among ROI-based measures, only medial femoral thickness showed a significant association at T_-2Y_, but it was less responsive than the CTh-Score (SRM=1.45 vs SRM<0.81 Supplementary Table 1).

### Associations with knee replacement outcomes

132 knees that subsequently underwent knee replacement were matched to 396 control knees (Table 4).

**Table 4.** Case–control comparison of cartilage morphology before knee replacement (KR) within the subsequent 2 years. Cross-sectional and longitudinal comparisons between knees that underwent KR during follow-up and matched control knees without KR. T_0_ is the last study visit with available MRI and radiographic data prior to surgery (within 2 years). T_−2Y_ and T_−4Y_ denote 2 and 4 years before T_0_, respectively. Values are median (IQR) for continuous variables and percentage for categorical variables. P values compare cases vs controls at each timepoint and for change scores (Δ). Measures: CTh-Score (0–100), medial minimum JSW (mm), and ROI-averaged cartilage thickness (mm) for femoral weight-bearing medial, femoral weight-bearing lateral, tibial medial, and tibial lateral compartments.

| T<sub>-4Y</sub> |  | Case<sub>KR</sub> | Control<sub>KR</sub> | p |
| --- | --- | --- | --- | --- |
|  | Number of knees | 132 | 396 |  |
|  | **Matched variables** |  |  |  |
|  | Age (y) | 65.0 (59.0;71.0) | 65.0 (59.0;71.0) | 0.99 |
|  | Sex (%F) | 62.10% | 62.10% | 1 |
|  | BMI (kg/m<sup>2</sup>) | 28.7 (26.1-31.6) | 28.4 (25.9-31.3) | 0.66 |
|  | KL0/1/2/3/4 (%) | 4.5/9.8/31.1/43.2/11.4 | 4.5/9.8/31.1/43.2/11.4 | 1 |
|  | **Non-matched variables** |  |  |  |
|  | JSW (mm) | 3.6 (2.1;4.8) | 3.6 (2.6;4.8) | 0.57 |
|  | Femur medial (mm) | 2.0 (1.7;2.4) | 2.1 (1.8;2.4) | 0.39 |
|  | Femur lateral (mm) | 2.4 (2.2;2.6) | 2.3 (2.2;2.5) | 0.22 |
|  | Tibia medial (mm) | 1.7 (1.6;1.9) | 1.7 (1.6;1.9) | 0.83 |
|  | Tibia lateral (mm) | 1.9 (1.7;2.2) | 1.9 (1.7;2.2) | 0.94 |
|  | CTh-Score (0-100) | 53.5 (38.5;71.5) | 46.4 (28.0;65.2) | **0.006** |
| T<sub>-2Y</sub> | JSW | 3.0 (1.6;4.7) | 3.4 (2.4;4.7) | 0.09 |
|  | Femur medial | 1.9 (1.6;2.2) | 2.1 (1.7;2.3) | 0.07 |
|  | Femur lateral | 2.4 (2.2;2.5) | 2.3 (2.1;2.5) | 0.34 |
|  | Tibia medial | 1.7 (1.6;1.9) | 1.7 (1.5;1.9) | 0.48 |
|  | Tibia lateral | 1.8 (1.6;2.2) | 1.9 (1.6;2.1) | 0.9 |
|  | CTh-Score | 64.2 (45.7;78.5) | 52.0 (32.77;70.3) | **<0.001** |
| T<sub>0</sub> | JSW | 2.3 (1.1;4.6) | 3.2 (2.0;4.6) | **0.01** |
|  | Femur medial | 1.8 (1.4;2.2) | 2.0 (1.7;2.3) | **0.005** |
|  | Femur lateral | 2.3 (2.1;2.5) | 2.3 (2.1;2.5) | 0.72 |
|  | Tibia medial | 1.6 (1.4;1.8) | 1.7 (1.5;1.9) | 0.09 |
|  | Tibia lateral | 1.9 (1.6;2.1) | 1.8 (1.6;2.1) | 0.58 |
|  | CTh-Score | 70.9 (52.4;84.0) | 57.0 (40.6;74.0) | **<0.001** |
| T<sub>-2Y</sub> - T<sub>-4Y</sub> | ΔJSW | -0.30 (-0.89;0.06) | -0.16 (-0.51;0.10) | **0.02** |
|  | ΔFemur medial | -0.08 (-0.25;0.02) | -0.06 (-0.17;0.03) | 0.07 |
|  | ΔFemur lateral | -0.02 (-0.09;0.05) | -0.01 (-0.09;0.04) | 0.96 |
|  | ΔTibia medial | -0.05 (-0.12;0.01) | -0.04 (-0.09;0.01) | 0.13 |
|  | ΔTibia lateral | -0.04 (-0.10;0.02) | -0.03 (-0.10;0.02) | 0.52 |
|  | ΔCTh-Score | 4.9 (0.15;11.7) | 3.7 (0.07;7.05) | **0.03** |
| T<sub>0</sub> - T<sub>-4Y</sub> | ΔJSW | -0.72 (-1.43; -0.10) | -0.30 (-0.78;0.00) | **<0.001** |
|  | ΔFemur medial | -0.09 (-0.39;0.00) | -0.07 (-0.20;0.03) | 0.01 |
|  | ΔFemur lateral | -0.01 (-0.09;0.04) | -0.03 (-0.12;0.04) | 0.27 |
|  | ΔTibia medial | -0.08 (-0.20; -0.01) | -0.05 (-0.13;0.01) | 0.01 |
|  | ΔTibia lateral | -0.09 (-0.17; -0.02) | -0.07 (-0.14; -0.01) | 0.1 |
|  | ΔCTh-Score | 9.4 (3.2-22.3) | 6.2 (2.2;12.7) | **0.006** |

At T_-4Y_, CTh-Scores were higher in knees that later underwent replacement compared with controls (53.5 [38.5; 71.5] vs. 46.4 [28.0; 65.2]; p = 0.006), despite no significant differences in JSW or regional cartilage thickness (all p ≥ 0.39). Differences in CTh-Score increased at T_-2Y_ (64.2 [45.7; 78.5] vs. 52.0 [32.8; 70.3]; p < 0.001) and were most pronounced at T_0_ (70.9 [52.4; 84.0] vs. 57.0 [40.6;74.0]; p < 0.001). At T_0_, JSW and medial femoral thickness regional thickness measures differ significantly between groups (p = 0.010 and p = 0.005).

Longitudinal changes in CTh-Score over the 2-year interval preceding T_-2Y_ significantly differ between groups (Δ 4.9 [0.15; 11.7] vs. 3.7 [0.07; 7.1]; p = 0.03). and also over the full 4-year interval, cases exhibited a larger increase in CTh-Score than controls (Δ 9.4 [3.2; 22.3] vs. 6.2 [2.2; 12.7]; p = 0.006). Changes in JSW were also significant, although CTh-Score showed slightly higher responsiveness over the 2-year interval and higher responsiveness over the full 4-year interval (SRM=0.86 vs SRM=0.76 Supplementary Table 1). Regional mean thickness changes showed only modest, inconsistent discrimination between groups (Supplementary Table 1).

In adjusted regression analyses (Table 2), the CTh-Score was independently associated with future knee replacement at both T_-4Y_ (OR per SD: 2.0 [1.4; 2.8], p < 0.001) and T_-2Y_ (1.89 [1.3; 2.9], p < 0.001). No ROI-based thickness measure demonstrated a significant independent association with knee replacement risk.

## Discussion

The CTh-Score, an automatic metric derived from high-resolution spatial patterns of cartilage thickness [5], outperformed conventional morphometric measures in detecting clinically meaningful clinical course milestones of OA. Across three matched case–control designs spanning incident radiographic OA, combined pain and structural progression, and knee replacement, the CTh-Score distinguished cases from controls as early as four years before the corresponding clinical milestone. It also remained independently associated with each outcome after adjustment for demographic factors and baseline radiographic severity and showed the most consistent longitudinal responsiveness compared with minimum joint space width (JSW) and region-based average cartilage thickness. To our knowledge, the CTh-Score is the first imaging-derived cartilage morphology metric to show consistent associations with these three major OA clinical course milestones across the disease spectrum.

A key finding of this study is the ability of the CTh-Score to detect cartilage damage as early as 4 years prior to radiographic onset, which is a capability not demonstrated by any conventional cartilage morphology metric to date. Knees that went on to develop radiographic OA already exhibited significantly higher CTh-Scores four years before diagnosis compared with those that did not, after controlling for demographics and radiographic severity. By contrast, neither regional average cartilage thickness nor minimum joint space width differentiated the two groups at that early time point. This observation is consistent with prior literature showing that conventional cartilage morphology measures, including semi-quantitative MRI gradings, fail to distinguish pre-radiographic cases from controls. In a nested case–control design similar to ours (the POMA study), semi-quantitative cartilage damage scores did not differ between incident OA cases and controls two years before disease onset [12]. Similarly, Reichenbach et al. [23], in a cross-sectional analysis of 948 participants from the Framingham Osteoarthritis Study, reported that neither cartilage volume nor regional thickness measurements were significantly lower in knees with Kellgren–Lawrence grades 1 or 2 compared with grade 0; differences only emerged at grades 3 and 4. The ability of the CTh-Score to identify pre-radiographic cartilage abnormalities is clinically significant because the silent phase of early OA (during which structural damage accumulates before radiographic signs manifest) represents a critical window of opportunity for disease-modifying interventions, before irreversible cartilage loss has occurred [24]. By shifting from regional averaging to whole-joint thickness pattern analysis, the CTh-Score enables the detection of meaningful inter-group differences years before they become apparent on conventional assessments.

The CTh-Score was also associated with four-year combined pain and structural progression in subjects matched for demographics, symptom intensity, and radiographic severity, including joint space width. Identifying structural and pain progression in OA is a major research priority, particularly for the design of clinical trials evaluating disease-modifying OA drugs (DMOADs). Because OA progresses slowly, enriching trials with patients at high risk of progression is essential to demonstrating treatment effects within feasible study durations. However, conventional metrics have proven inadequate for this purpose. In the IMI-APPROACH study, for instance, patient selection was guided by machine learning algorithms incorporating demographics, questionnaire responses, and radiographic variables including joint space width to predict structural or pain progression [25]. While the algorithm’s predictions aligned well with observed pain progression, this was not the case for structural progression [26]. One plausible explanation is that inter-individual variability in baseline conventional cartilage thickness measures makes them poor predictors of future structural change. Our findings support this interpretation. Regional mean cartilage thickness did not differ between progressors and controls either four or two years before radiographic progression was documented. Previous work [27] in the same cohort showed that cartilage loss over 24 months, defined as the change in cartilage thickness between two measurements taken 24 months apart, was associated with subsequent progression, which is consistent with our results. In contrast, the CTh-Score, which is designed to provide a severity metric that can be compared directly across individuals from a single measurement, showed significant differences between groups at both timepoints. These results are further corroborated by semi-quantitative MOAKS analyses of the FNIH cohort, which similarly identified differences in cartilage damage between progressor and non-progressor groups [13].

The structural progression criterion adopted in the FNIH is minimum joint space width [22], and the most responsive measurements are obtained from the medial compartment, with lateral-predominant progressors excluded from the analysis. Within this medial-focused context, changes in medial regional cartilage thickness averages were indeed associated with progression status and differed between groups. Notably, the CTh-Score also demonstrated significant associations with progression despite not being a compartment-specific measure, and it exhibited superior responsiveness compared with regional averages.

Knee replacement represents a definitive clinical endpoint for end-stage OA. Previous studies using the OAI have reported that semi-quantitative MRI-based cartilage damage was significantly greater in knees destined for replacement compared with controls 12 months before surgery [14]. Our findings with the CTh-Score align with and extend these observations, demonstrating significant differences between future knee replacement cases and controls at T_−4Y_ (approximately six years before surgery) and T_−2Y_ (approximately four years before surgery). This analysis is inherently challenging because knee replacement decisions are influenced by factors beyond structural severity alone, including patients’ preferences, availability of insurance, comorbidities, orthopedic surgeons’ decision making and access to surgical care: all factors that may contribute to structurally and symptomatically eligible patients not undergoing the procedure. Despite these confounders, the CTh-Score demonstrated the highest responsiveness among all evaluated metrics and was the only cartilage measure independently associated with future knee replacement after adjustment for covariates.

This study has limitations that should be acknowledged. The matched case–control design, while effective for evaluating associations across the OA continuum, is retrospective in nature and does not take into account unmeasured confounding. Validation of these findings in appropriately designed prospective studies and independent longitudinal cohorts is warranted. Future work should also evaluate the utility of the CTh-Score in additional clinical settings, including clinical trial enrichment strategies and treatment response monitoring. We encourage the broader research community to leverage the publicly available repository of CTh-Maps and CTh-Scores for alternative study designs within the OAI.

In conclusion, the CTh-Score automatically derived from high-resolution cartilage thickness maps, detects clinically meaningful OA milestones with greater sensitivity than conventional averaged morphometric measures. By analyzing spatial patterns across the entire joint surface rather than regional averages, this approach addresses fundamental limitations of current conventional cartilage assessment methods. Together, these findings support the utility of CTh-Maps as informative representations of disease-specific structural fingerprints in cartilage, and of the CTh-Score as an effective metric of OA cartilage severity across different clinically relevant milestones in the clinical course of knee OA.

**Figure 1.**
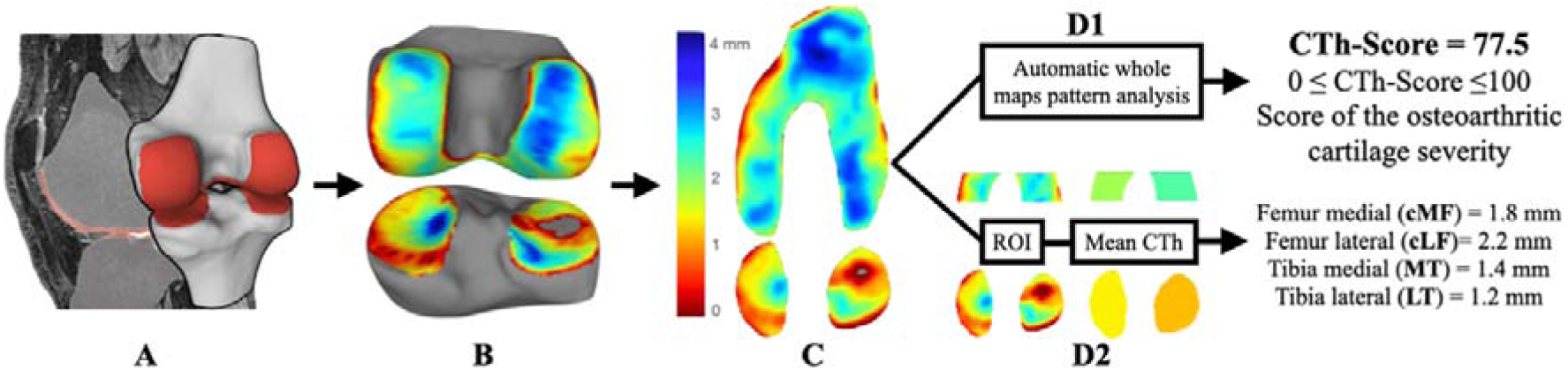
Automated MRI-based assessment of cartilage morphology and severity. Workflow for generating anatomically standardized cartilage thickness maps (CTh-Maps) and the Cartilage Thickness Score (CTh-Score) from knee MRI. (A) Automated cartilage segmentation from sagittal DESS images. (B) Three-dimensional anatomical standardization to establish spatial correspondence of femorotibial surfaces among and within participants. (C) Two-dimensional projection to produce high-resolution standardized CTh-Maps. (D1) Deep learning–based pattern recognition applied to CTh-Maps to derive the CTh-Score (0–100), representing a continuous cartilage severity score from healthy to end-stage osteoarthritis. (D2) Conventional region-of-interest (ROI) analysis: predefined ROI cropping and averaging to obtain compartment-level mean cartilage thickness.

**Figure 2.**
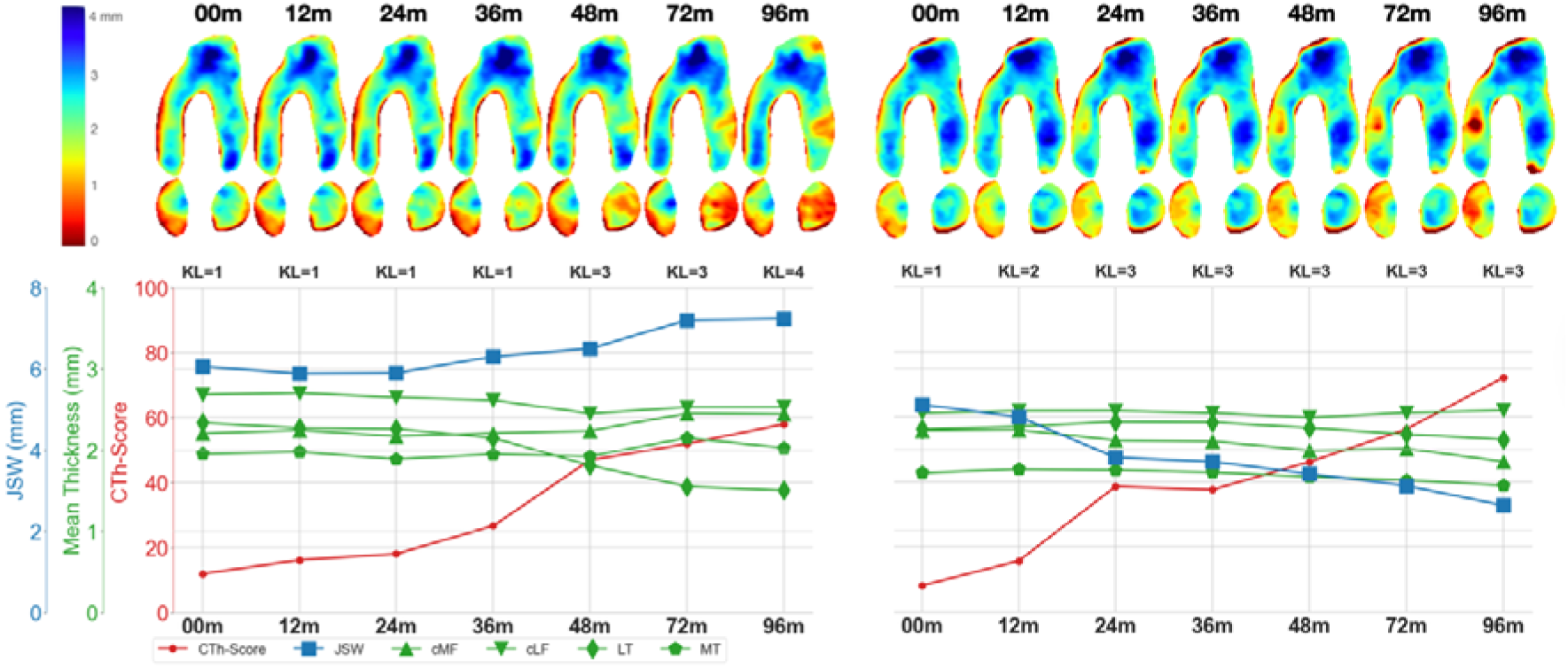
Example longitudinal cartilage thickness trajectories in two OAI knee participants with distinct phenotypes. Representative evolution of cartilage damage patterns for two independent OAI knee participants illustrating medial- versus lateral-predominant disease. Subject A shows predominantly lateral cartilage loss, whereas Subject B shows predominantly medial cartilage loss. Panels display radiographic minimum joint space width (JSW; medial compartment), ROI-averaged cartilage thickness (mm)(cMF: central medial femur; cLF: central lateral femur; MT: medial tibia; LT: lateral tibia), and CTh-Score derived from CTh-Maps. Notably, medial JSW primarily reflects medial compartment narrowing and may not capture lateral-dominant disease. The CTh-Score is computed without using timepoint information and provides a temporally consistent severity signal that tracks lesion progression across compartments.

## Data Availability

All data produced in the present study are available upon reasonable request to the authors

---

**Supplementary Table 1.** Responsiveness of cartilage morphology metrics across osteoarthritis milestones. Responsiveness of CTh-Score, medial minimum JSW, and ROI-averaged cartilage thickness (cMF, cLF, MT, LT) across the three matched case–control designs: radiographic onset, combined pain and structural progression, and knee replacement. Responsiveness is summarized over 4-year (T_−4Y_→T_0_) and 2-year (T_−4Y_→T_−2Y_) intervals relative to the matching timepoint. Standardized response mean (SRM) is the absolute mean change / SD of change. SRM of JSW in Progression is not applicable (na) because it is used to define the structural progression. Abbreviations: JSW, joint space width; cMF/cLF, central weight-bearing femoral ROIs; MT/LT, tibial ROIs.

|  |  | Radiographic onset<br>n=304 | Progression<br>n=164 | Knee replacement<br>n=132 |
| --- | --- | --- | --- | --- |
| T<sub>-4Y</sub>-T<sub>-2Y</sub> | JSW | 0.31 | na | 0.56 |
|  | Femur medial | 0.16 | 0.62 | 0.31 |
|  | Femur lateral | 0.02 | 0.09 | 0.23 |
|  | Tibia medial | 0.20 | 0.55 | 0.35 |
|  | Tibia lateral | 0.37 | 0.32 | 0.42 |
|  | CTh-Score | **0.86** | **0.97** | **0.57** |
| T<sub>-4Y</sub>-T<sub>0</sub> | JSW | 0.71 | na | 0.76 |
|  | Femur medial | 0.36 | 0.81 | 0.56 |
|  | Femur lateral | 0.16 | 0.15 | 0.35 |
|  | Tibia medial | 0.28 | 0.81 | 0.56 |
|  | Tibia lateral | 0.63 | 0.67 | 0.70 |
|  | CTh-Score | **1.41** | **1.45** | **0.86** |

